# Clinical outcomes and survival of individuals with methylmalonic acidemia, propionic acidemia, classic homocystinuria, and remethylation disorders identified through newborn screening

**DOI:** 10.1101/2023.09.15.23295546

**Authors:** Anna T. Reischl-Hajiabadi, Elena Schnabel, Florian Gleich, Katharina Mengler, Martin Lindner, Peter Burgard, Roland Posset, Svenja Lommer-Steinhoff, Sarah C. Grünert, Eva Thimm, Peter Freisinger, Julia B. Hennermann, Johannes Krämer, Gwendolyn Gramer, Dominic Lenz, Stine Christ, Friederike Hörster, Georg F. Hoffmann, Sven F. Garbade, Stefan Kölker, Ulrike Mütze

**Affiliations:** Division of Child Neurology and Metabolic Medicine, Center for Child and Adolescent Medicine, University Hospital Heidelberg, Heidelberg, Germany; University Children’s Hospital, Frankfurt am Main, Germany; Department of General Pediatrics, Adolescent Medicine and Neonatology, Medical Center, University of Freiburg, Faculty of Medicine, Freiburg, Germany; Department of General Pediatrics, Neonatology, and Pediatric Cardiology, University Children’s Hospital, Heinrich Heine University Düsseldorf, Düsseldorf, Germany; Children’s Hospital Reutlingen, Klinikum am Steinenberg Reutlingen, Reutlingen, Germany; Villa Metabolica, Department of Pediatric and Adolescent Medicine, University Medical Center Mainz, Mainz, Germany; Department of Pediatric and Adolescent Medicine, Medical School, Ulm University, Ulm, Germany; Department for Inborn Metabolic Diseases, University Children’s Hospital, University Medical Center Hamburg-Eppendorf, Hamburg, Germany

**Keywords:** neonatal screening, methylmalonic aciduria, propionic aciduria, MTHFR deficiency, cblC deficiency, neonatal vitamin B_12_ deficiency

## Abstract

The current German newborn screening (NBS) panel includes 13 inherited metabolic diseases (IMDs). In addition, the NBS pilot study in Southwest Germany identifies individuals with methylmalonic acidemia (MMA), propionic acidemia (PA), cystathionine β-synthase (CBS) deficiency, remethylation disorders [e.g. cobalamin (cbl) C and methylenetetrahydrofolate reductase (MTHFR) deficiency], and neonatal cbl deficiency through a combined second-tier algorithm. The long-term health benefits of screened individuals are evaluated in a prospective multicenter observational study.

Twenty-seven individuals with IMDs [MMA (N=6), PA (N=13), cblC deficiency (N=5), MTHFR deficiency (N=2) and CBS deficiency (N=1)] and 42 with neonatal cbl deficiency were identified by the NBS pilot study and followed for a median of 3.6 years. Seventeen IMD patients (63%) experienced at least one metabolic decompensation, 14 of them neonatally and six even before the NBS report (cbl-nonresponsive MMA, PA). Three PA patients died despite NBS and immediate treatment. Fifteen individuals (79%) with MMA or PA and all with cblC deficiency presented with permanent, mostly neurological symptoms, while individuals with CBS, MTHFR and neonatal cbl deficiency had a favorable outcome.

Utilizing a combined second-tier algorithm we demonstrate that NBS and specialized metabolic care result in substantial benefits for individuals with CBS deficiency, MTHFR deficiency, neonatal cbl deficiency, and to some extent, cblC deficiency and cbl-responsive MMA. However, its advantage is less evident for individuals with cbl-nonresponsive MMA and PA.

## Introduction

Newborn screening (NBS), performed in the first days of life (1), aims to identify affected individuals in the pre-clinical, or at least early disease state to prevent irreversible damage and untimely death (2). In the last 60 years, NBS programs have been successfully established in many countries worldwide. Although agreeing on the same set of NBS criteria, national NBS panels vary widely (3). Since it might be difficult to meet all NBS criteria for rare diseases that are considered candidates for extended NBS panels, pilot studies demonstrating the feasibility, diagnostic process quality, and patient benefit are an important tool to evaluate these candidate diseases through collection of structured data and careful evaluation (4). Since 2016, the NBS center at the University Hospital Heidelberg has been coordinating a pilot panel utilizing a combined second-tier algorithm for identification of individuals with different types of methylmalonic acidemia (MMA), propionic acidemia (PA; MIM #606054), cystathionine-β-synthase (CBS; EC no. 4.2.1.22) deficiency (MIM #236200), remethylation disorders (#236250, #277400, #277410, #236270, #250940, #277380, #309541, #614857), or neonatal cobalamin (cbl; synonym, vitamin B_12_) deficiency (5–8).

The phenotypic spectrum of isolated MMA is wide, depending on the residual enzyme activity of methylmalonyl-CoA mutase (mut, EC no. 5.4.99.2) and the responsiveness to its cofactor 5’-adenosyl-cbl. Cbl-nonresponsive individuals (mut^0/-^-type; MIM #251000; and cblB-type MMA; MIM #251110) usually present with a severe early-onset form which is characterized by life-threatening metabolic decompensations already during the neonatal period, while the disease severity is more attenuated in individuals with cbl-responsive MMA (cblA-type; MIM #251100) (9). Furthermore, responsiveness to cbl is associated with increased life expectancy and lower disease severity compared to cbl-nonresponsive individuals (10–12). Since about two thirds of individuals with MMA remained asymptomatic until day 8 in a previous European cohort study (13), NBS might be an effective tool to prevent neonatal disease manifestation and to improve clinical outcome in a relevant number of screened individuals, particularly for individuals with cbl-responsive and late-onset forms of MMA (13). Similarly to MMA, individuals with early-onset PA present with life-threatening neonatal metabolic decompensations, and those with late-onset PA manifest at a median age of 5-6 months (9). It was estimated that NBS could identify about 50% of individuals with PA pre-symptomatically (13,14). However, the long-term clinical benefit of NBS for individuals with PA has not yet been convincingly shown (14).

Depending on the degree of responsiveness to pyridoxine (synonym, vitamin B_6_) in CBS deficiency, first clinical symptoms occur in early childhood (15–17) at a median age of 4 years (18) in the nonresponsive group, while individuals with responsive forms (17) become symptomatic in the second (full responders) or even third (extreme responders) decade (18). Due to this clinical latency, NBS allows pre-clinical identification and treatment, leading to the prevention of ocular and vascular manifestations and expectation of nearly normal development (15, 18, 19). However, individuals with full or extreme pyridoxine responsiveness are considered to be only partially (7) or even not (15) identified by NBS.

For remethylation disorders, early therapeutic intervention in individuals with methylenetetrahydrofolate reductase (MTHFR; EC no. 1.5.1.20) deficiency enables significant clinical improvement across all disease severities (19–21). In analogy, NBS for cobalamin C (cblC) deficiency is also thought to be beneficial. The initiation of early treatment with hydroxo-cbl, betaine, folic or folinic acid, and carnitine leads to better developmental outcomes (22), reduced premature death, reduced frequency and severity of hematological manifestations, less failure to thrive and neurodevelopmental impairment compared to symptomatically diagnosed individuals (19).

MMA, PA, CBS deficiency, and remethylation disorders are not included in the regular German NBS yet (1). The present pilot study aims at evaluating the health benefits of screened and early treated individuals with these IMDs.

## Material and Methods

### Study design

In 2016, a multicenter NBS pilot study in Southwest Germany has been initiated evaluating 28 candidate diseases for an extended national NBS panel in Germany (NGS2025; Trial registration ID: DRKS00025324; PI: G. F. Hoffmann (23)). The study has been approved by the ethics committee of Heidelberg University Hospital (No S-533/2015). Data on outcome of individuals identified by this NBS pilot study is collected in a prospective multicenter observational study that was implemented in 2005 (NGS2020 and NGS2025; trial registration: DRKS00013329; PI: S. Kölker; (24)). This study has been approved by the local ethics committee of Heidelberg University Hospital (No. S-104/2005) and consecutively by all participating study sites (Freiburg, Düsseldorf, Mainz, Reutlingen and Ulm, all Germany). Inclusion criteria are (A) date of birth on or after January 1, 1999; (B) confirmation of the suspected IMD identified by NBS (national NBS program and pilot panels) according to the national guidelines and international recommendations by using additional metabolite, enzyme and/or genetic tests; and (C) written informed consent before enrollment (24).

### Study population

Individuals enrolled in the observational study were followed according to a standardized protocol. Comprehensive regular follow-up information was obtained at least at one time point at the ages of 1.5 ± 0.5 years, 3.5 ± 0.5 years, 5.5 ± 0.5 years, 9 ± 1 years, 14 ± 1 years or 18 ± 1 years by structured clinical examination, recording of medical history, and neuropsychological testing. The latter was performed by Denver Developmental Screening Test (DDST) at the age of 1.5 ± 0.5 years and IQ-based tests in older individuals [i.e. Wechsler Preschool and Primary Scale of Intelligence (WPPSI-III, WPPSI-IV), Wechsler Intelligence Scale for Children (WISC-IV, WISC-V), and Wechsler Adult Intelligence Scale (WAIS-IV)]. Testing with DDST was continued in children with severe developmental delay in whom IQ-based testing was not possible. Screened individuals with a confirmed diagnosis who died before the first visit were included in the analysis with an anonymized basic data set. For patients younger than one year of age at data collection, preliminary study visits were recorded and marked data were included in the analysis. Cut-off for data analysis was January 31^st^, 2023.

### Classification and definitions

All individuals with any of the NBS candidate diseases included in this study have a risk for lifelong metabolic decompensations, except for neonatal cbl deficiency. *Metabolic decompensation* was defined as any event that required hospitalization (at least one night) for a subclinical biochemical derangement (i.e. hyperammonemia, hypoglycemia, metabolic acidosis, and/or dehydration) or a clinical presentation (i.e. altered consciousness, thromboembolic event, and/or an acute neurologic event indicative of metabolic decompensation). Metabolic decompensations were classified as *early onset* (EO; <29 days of life) or *late onset* (LO; >28 days of life). Following literature search and evaluation, a set of *disease-specific symptoms* was defined (Supplementary Table) (11, 15, 21). At each study visit, individuals were examined for disease-specific symptoms. Patients were classified as *symptomatic* if individual was stated symptomatic at the time of the first NBS report and subsequent start of treatment or if the patient was recorded symptomatic at any study visit point and/or if at least one metabolic decompensation occurred. In order to evaluate management and care, adherence to therapy recommendations (T) and biochemical checkups as well as clinical monitoring for comorbidities (M) (11, 15, 21) were classified as follows: T3: therapy according to the guideline, T2: therapy in partial according to the guideline, T1: no therapy; M3: all recommended biochemical and clinical monitoring, M2: isolated biochemical monitoring, M1: no monitoring. From this, the following categories were defined for the analysis: 1 (T3+M3), 2 (T3+M2), 3 (T2+M3), 4 (T2+M2) and 5 (T1+M1 or T1+M2 or T2+M1). *Migration background* was defined as at least one parent born with non-German citizenship.

### Statistical analysis

Statistical analyses were performed using IBM SPSS® 29 and R language for statistical computing version 4.3.1. For continuous variables, the mean, median, standard deviation, and interquartile range were calculated. For categorical variables, the absolute and relative frequencies were calculated.

For the calculation of the hospitalization rate of the normal population in Germany, data from the Federal Statistical Office (DESTATIS) on birth and death rates (26, 27) and data from the German Society for Pediatrics and Adolescent Medicine (DGKJ) (28) were used. From these data, the annual hospitalization rate per child was estimated as 0.08 and the total pediatric hospitalization rate 1.4 times (≤18 years of life).

Data which could not be retrieved and implausible data that could not be verified were treated as missing values for the respective analysis.

## Results

### Study cohort

From January 1999 to September 2022, 183 infants with MMA (N=10), PA (N=14), CBS deficiency (N=1), remethylation disorders (N=9), or neonatal cbl deficiency (N=149) were identified by the NBS pilot study. Sixty-nine of them participated in the outcome study (Table 1; Table 2), including MMA (N=4 mut^0^-type; N=2 cblA-type MMA), PA (N=13), CBS deficiency (N=1), remethylation disorders (N=5 cblC deficiency; N=2 MTHFR deficiency) and neonatal cbl deficiency (N=42). Nearly all participants were born at term (median 39 weeks of gestation, range 36-41). Migration background was reported in 48% and known consanguinity in 33%. The median age of follow-up was 3.6 (range 0-17.8) years. The short-term outcome of a subgroup of individuals with neonatal cbl deficiency has been reported previously (N=32 (29)). This group is evaluated and displayed separately from the IMDs (Table 2).

**Table 1:**
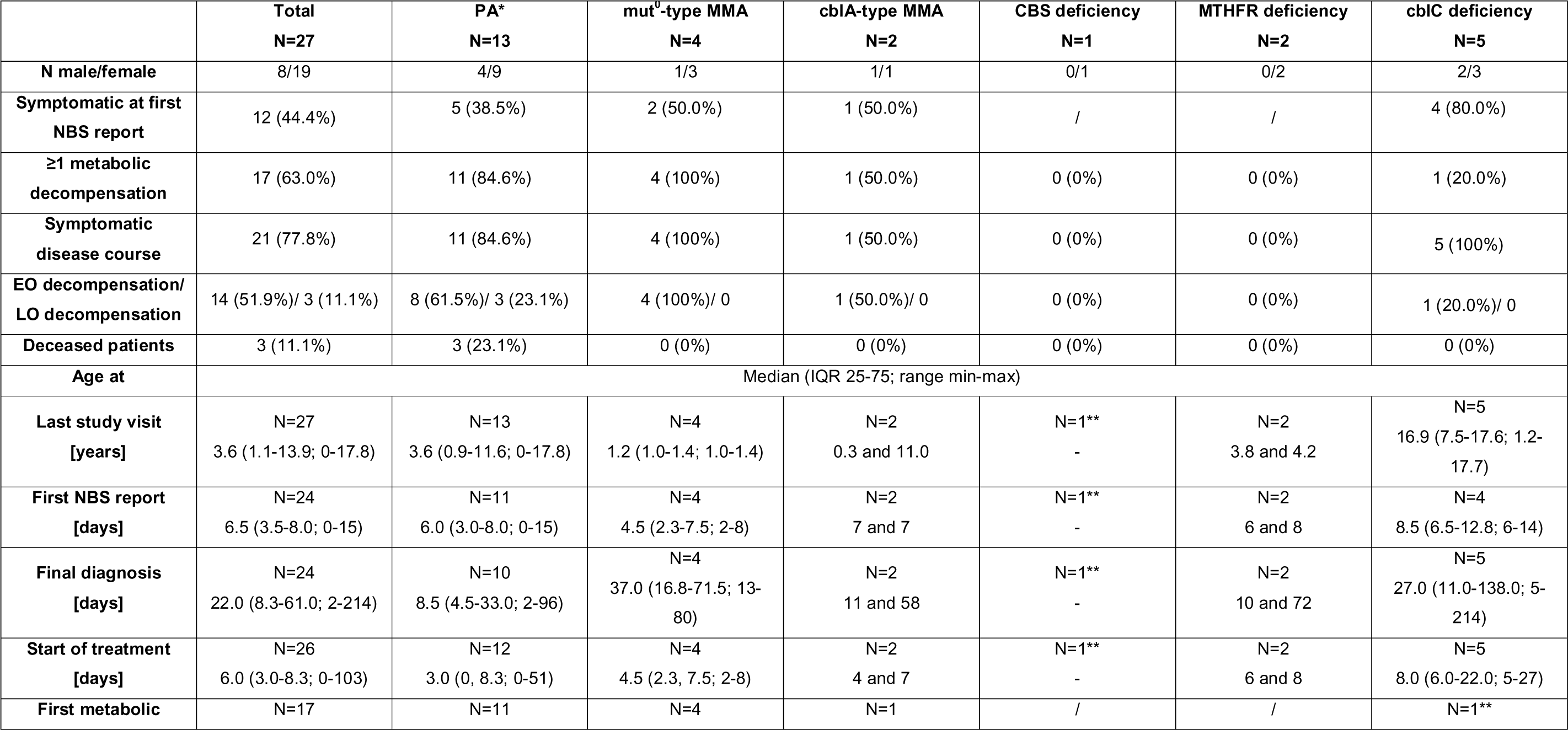
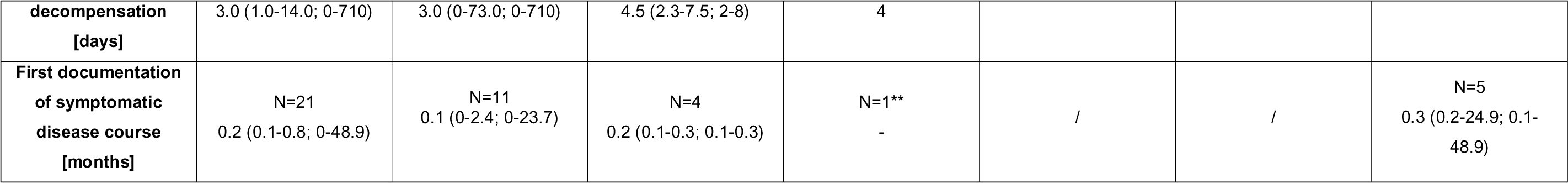
Patients’ characteristics of screened individuals with PA, MMA, CBS deficiency and remethylation defects. Data on the neonatal cobalamin deficiency cohort are displayed separately (Table 2). *One patient with an additional diagnosis of thiamine-responsive encephalopathy. ** To avoid to publish identifying data, ages are not given for N=1, however data is included in the total column.

**Table 2:** Neonatal cobalamin deficiency.

|  |  |
| --- | --- |
| **N (male/female)** | 42 (22/20) |
| **Age appropriate result of DDST** | N=34<br>34 (100%)<br>Median age 1.17 (1.08, 1.50; 0.83-1.92) years |
|  | **Median (IQR; Range)** |
| **Age at last study visit [years]** | N=42<br>1.1 (0.8, 1.4; 0.1-1.9) |
| **Age at first NBS report [days]** | N=41<br>9.0 (7.5, 15.0; 0-68) |
| **Age at final diagnosis [days]** | N=40<br>23.5 (18.0, 31.8; 6-116) |
| **Age at start of treatment [days]** | N=39<br>25.0 (17.0, 31.0; 6-116) |

### Survival

Three patients (all PA) died before the first study visit (age range first week to 2 years). This results in a mortality rate of 23% for PA identified by NBS (Table 1).

### Metabolic decompensation despite NBS

Two thirds of screened patients with a lifelong risk of metabolic decompensations (N=17/27) experienced at least one decompensation, 14 of them (82%) already neonatally (EO) at a median age of 3 (range 0-20) days (Figure 1, Table 1), among them 10 patients (59%) who presented before the 5^th^ day of life and 12 patients (86%) before the 7^th^ day of life.

**Figure 1:**
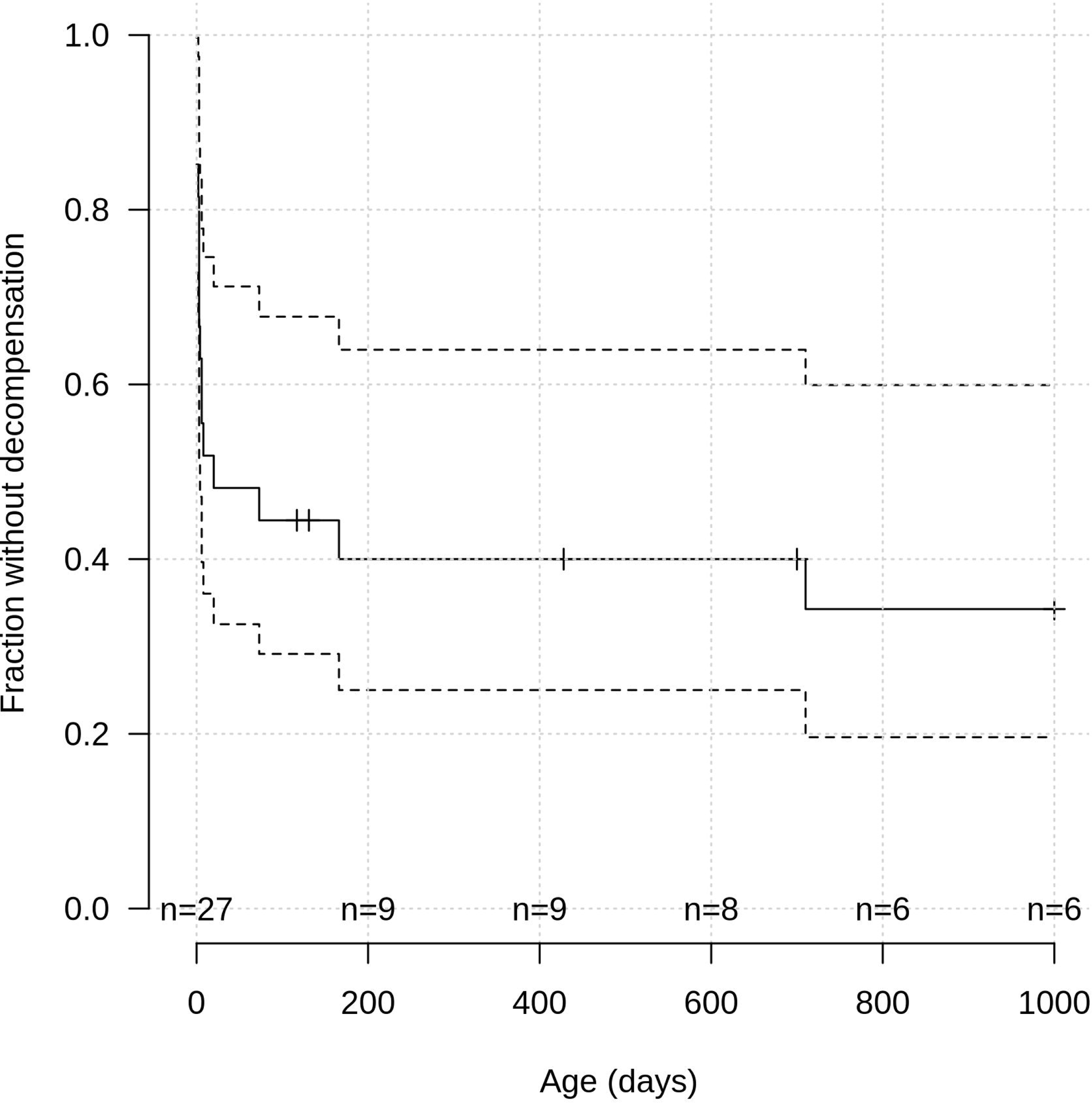
Kaplan-Meier analysis with 95% confidence interval (dotted lines) of the age at first metabolic decompensation for screened individuals with an IMD included by the pilot study [PA, MMA, CBS deficiency, and remethylation disorders (N=17)]. Numbers of followed patients are specified for age.

Of note, EO metabolic decompensation clinically occurred in 6 (43%) newborns before the NBS result (N=4 PA; N=1 cblA-type MMA; N=1 cblC deficiency) and in 7 (50%) after the NBS result (N=3 PA; N=4 mut^0^-type MMA) was reported. However, in four of the latter (N=1 PA; N=3 mut^0^-type MMA) first signs of metabolic decompensation were found in the initial laboratory analysis done for confirmation. For one infant with PA who presented with a metabolic decompensation already at the first day of life, exact time of NBS report was not recorded.

In three children with PA (18%), the initial metabolic decompensation occurred after the neonatal period (LO) at a median age of 5.5 (range 2.4-23.7) months during an episode of catabolic stress.

Overall reported outcome (N=14) after the initial metabolic decompensation was favorable in 11 patients (79%) without changes to the clinical status before; however, one PA patient died in the first metabolic decompensation and two children (PA) survived with neurological residuals (muscular hypotonia; movement disorder).

In total, 118 metabolic decompensations were documented within the first 6 years of life. Most of them occurred in children with PA up to the age of three years (Figure 2).

**Figure 2:**
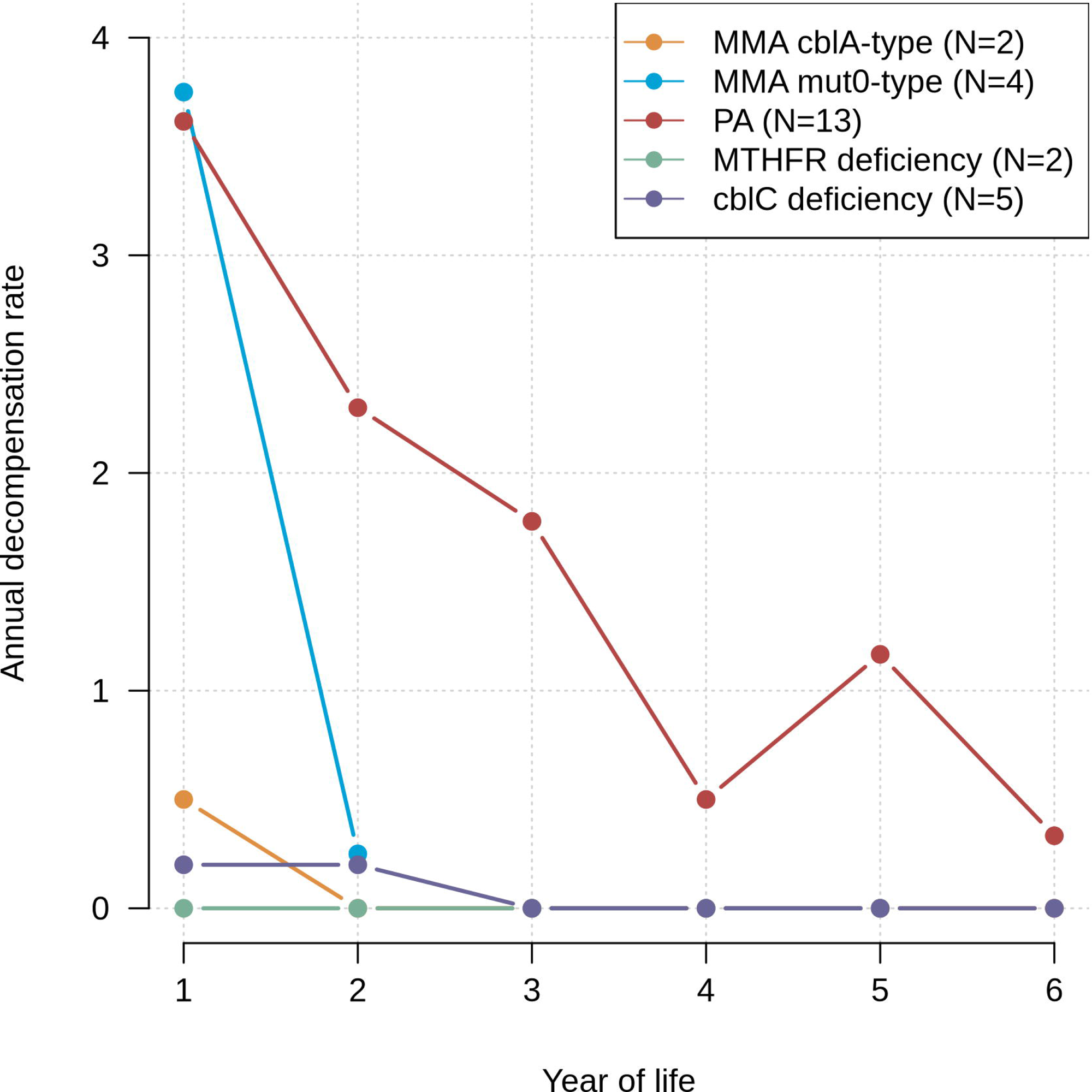
Disease-specific number of annual metabolic decompensations (N). The single patient with CBS deficiency had no decompensation and is therefore omitted from the figure.

### High hospitalization rates for PA, MMA, and cblC deficiency

Except for one patient with MTHFR deficiency, all patients were admitted to hospital at least once. During the first six years, mean number of hospitalizations per year ranged from 0.5 to 5.3 for the different IMDs (Figure 3), with the highest rates for PA, MMA and cblC deficiency and during the first year of life.

**Figure 3:**
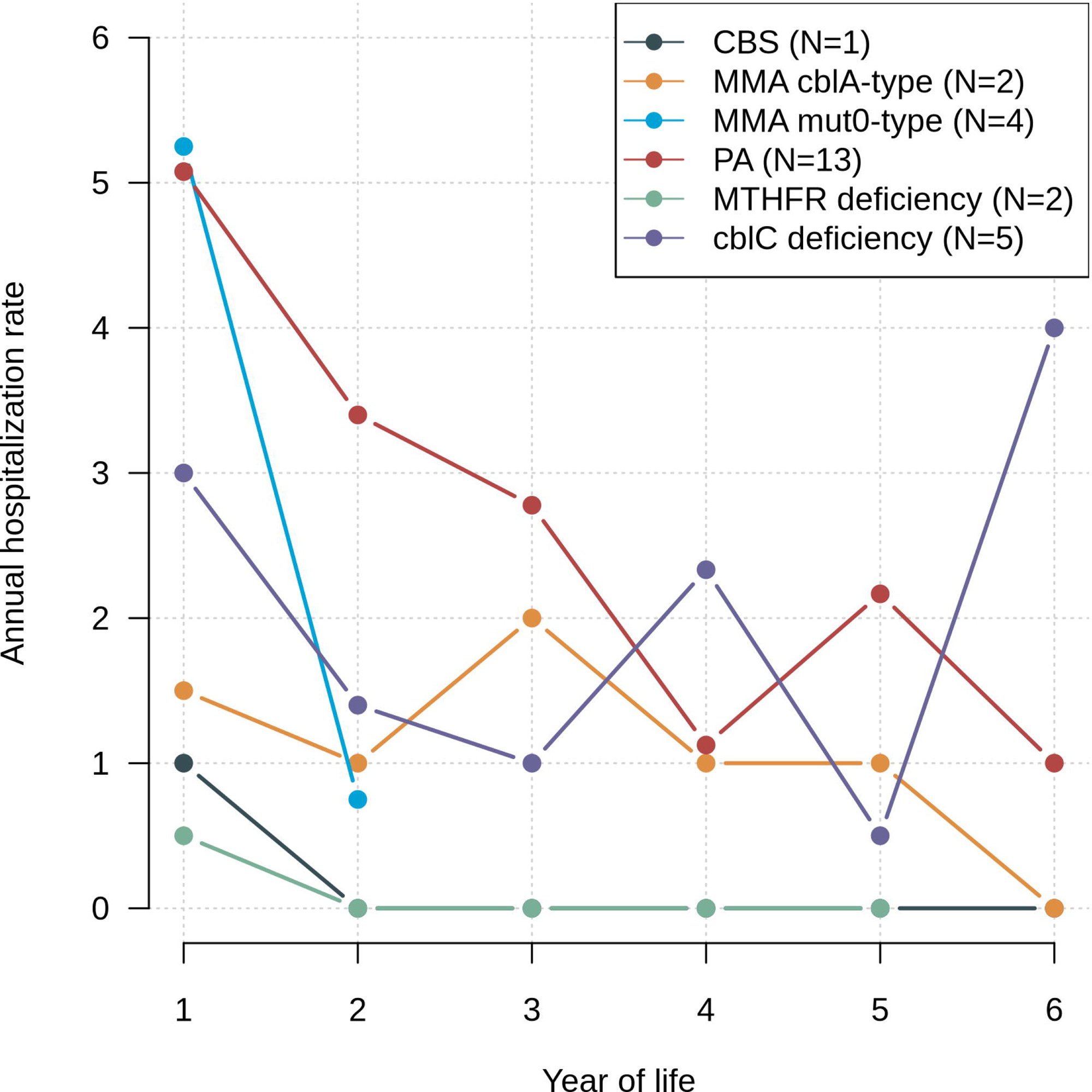
Disease-specific number of annual hospitalizations (N) of study patients.

By the age of six years, a total of 229 hospitalizations [median duration 4 (range 0-57) days] were reported for the 27 children with screened IMDs of our cohort. That revealed an overall mean number of hospitalizations of 8.5 in the first six years of life, i.e. a mean number of 1.4 hospitalizations per year, which markedly exceeds the mean number of pediatric hospitalizations until the 18^th^ birthday; i.e. 1.4 times (26–28).

Reasons for admission (multiple answers possible) were predominantly (47%) imminent metabolic decompensations (98% of the hospitalizations in children with PA and MMA), infectious diseases (40%), and nutritional problems (34%), while scheduled visits (21%) and new neurologic symptoms (e.g. seizures 4%) were less frequently reported as reason for admission.

### Symptomatic disease course despite NBS and early treatment

Sixteen participants (59%) presented with disease-related symptoms at the last study visit (Table 1, Supplementary Table), most of them showed neurological symptoms (multiple answers possible), especially impaired fine and gross motor function (41%), impaired speech (41%), and/or cognition (44%). Children with PA developed cardiac symptoms: cardiomyopathy (N=1, first report in early childhood) and prolonged QT_c_ interval [N=3, median age 9.8 (range 8.3-17.5) years]. Renal disease manifestation was not observed for MMA patients until last follow up at a median age of 1.2 (mut^0^-type MMA) and 5.7 (cblA-type MMA) years, respectively. Optic nerve atrophy was detected in three cblC deficiency patients, first at a median age of 7 (range 4-7.6) years.

Overall, considering metabolic decompensations and disease-related symptoms, 21 children of the study cohort (78%) experienced a symptomatic disease course until the last visit at a median age of 3.6 years (Table 1). Noteworthy, all individuals with mut^0^-type MMA and cblC deficiency became symptomatic.

### Impaired cognitive outcome and participation in regular schooling despite NBS and early treatment

DDST and/or IQ test results were available for 17 patients of the IMD cohort. DDST was performed in 11 children at a median age of 1.9 (range 1-16.3) years. Four of them (36%) obtained age-appropriate results (N=2 cblC deficiency and each one with mut^0^-type MMA and MTHFR deficiency), while in seven of them (64%) DDST was not considered appropriate for age. In detail, development was delayed globally (N=1 PA, N=2 cblC deficiency) or in several subdomains (two children each with mut^0^-type MMA and PA; multiple counts possible): 75% each for fine and gross motor skills, and speech, and 50% for social contact. Last IQ-based tests [N=6; performed at median age 10.0 (range 3.3-20.9) years] revealed an overall mean (SD) IQ of 91.7 (11.1), ranging from one patient with cblC deficiency (IQ 79), four PA patients [90.5 (6.6)] to one patient with cblA-type MMA (IQ 109).

Evaluating kindergarten and school attendance, we found about half of the children, 3 years and older (N=7/15), to attend kindergarten (N=4 PA; N=1 cblA-type MMA; N=2 MTHFR deficiency), two of them (both PA) requiring assistance. Ten children already reached school age (six years), only three of them attended a regular school (one each with PA, cblC deficiency, and cblA-type MMA), another one did with assistance (cblC deficiency), while more than half of the cohort (N=3 PA; N=2 cblC deficiency) attended schools for children with special needs, and/or mentally or physically disabled children. Information about school placement was missing for one child (PA).

### Treatment and monitoring following existing guidelines

Until the last study visit, all IMD patients (N=27) received metabolic treatment according to current guidelines and recommendations (11, 15, 21) (i.e. diet and/or medication). In 24 of them (89%), biochemical monitoring and screening for comorbidities was performed according to guidelines, while in three patients (11%) monitoring was primarily based on biochemical parameters. None of the children in our cohort received an organ transplant during the observation period.

Subsequent to the developmental delay and cognitive impairment, children in this study required more often allied health professionals (speech therapy, occupational therapy, and physiotherapy) compared to the reference population (Table 3) (30).

**Table 3:** Use of allied health professionals. documented for study participants with PA, MMA, CBS deficiency and remethylation defects (N=17; NGS2025 cohort) at study visits at the ages of 1.5 and 3. 5 years (N=25) and 5.5 and 9 years (N=9) and compared to data obtained from AOK, Germany’s largest health insurance company (30)

|  | Speech therapy<br>[%] | Occupational therapy<br>[%] | Physiotherapy<br>[%] |
| --- | --- | --- | --- |
| **NGS2025 cohort - Study visits 1.5 and 3.5 years** | **20** | **4** | **28** |
| **AOK 0-4 years** | **2** | **0** | **5** |
| **NGS2025 cohort - Study visits 5.5 and 9 years** | **44** | **33** | **33** |
| **AOK 5-9 years** | **13** | **7** | **3** |

\*Results of patients with >1 study visit were aggregated to avoid bias.

### Neonatal cobalamin deficiency

Forty-two individuals with neonatal cbl deficiency who were identified by the combined second-tier algorithm participated in the outcome study. Until last follow-up for clinical and cognitive outcome at a median age of 1.1 (range 0.1-1.9) years (Table 2), all of them were age appropriate according to DDST and none of them developed disease-related symptoms after adequate treatment.

## Discussion

The overall aim of NBS programs is to prevent irreversible harm and to improve clinical outcomes and survival of screened individuals with an NBS target disease through early and correct identification and initiation of treatment. With the introduction of new techniques for population-based screening, NBS disease panels are steadily but still slowly growing. To evaluate the potential health-related benefits from NBS using a combined screening algorithm (5–7), 27 screened individuals with MMA, PA, CBS deficiency, remethylation disorders (cblC deficiency, MTHFR deficiency), and 42 individuals neonatal cbl deficiency were carefully followed until a median age of 3.6 years.

Despite NBS and early treatment according to the current guidelines, the clinical outcomes of individuals with cbl-nonresponsive MMA, PA and cblC deficiency were less favorable compared to those with cbl-responsive MMA, CBS deficiency, MTHFR deficiency, and neonatal cbl deficiency: (1) Survival was favorable (100%) for screened individuals with MMA, CBS deficiency and remethylation disorders, while premature death was still high in screened PA patients (23%). (2) Two thirds of screened individuals with IMDs (all MMA, PA, and cblC deficiency) presented with at least one episode of metabolic decompensation, the majority (82%) already neonatally and nearly half of them (43%) even before the NBS result became available. (3) By the last study visit (median, 3.6 years) three quarters of the screened individuals with IMDs became symptomatic; 84% of the children with PA and MMA and all with cblC deficiency. (4) In contrast, individuals with CBS and MTHFR deficiency remained asymptomatic. (5) A favorable outcome for individuals with neonatal cbl deficiency was clearly demonstrated, confirming the previous study (29).

### Impact of NBS and early treatment on mortality and morbidity

All individuals with screened MMA of the present cohort survived, while natural history data reported high mortality rates for mut^0^-type MMA patients (41%) with a median age of death of 2 years (range 2 days to 22.6 years), but a much lower mortality rate for cblA-type MMA (5%) (10). This points to a potential health benefit of NBS and early treatment for mut^0^-type MMA. Individuals with mut^0^-type MMA usually present with an neonatal metabolic decompensation, beginning at a median age of 5 days (10). The present study confirmed this very early age at onset (4.5 days) for mut^0^-type MMA. For individuals with cblA-type MMA, a later onset of symptoms (median, 25 days) was reported previously (10, 12). In this cohort, one patient with cblA-type MMA experienced an EO decompensation prior to the NBS result but had a normal IQ and did not develop further symptoms on treatment, while the second patient remained asymptomatic after early diagnosis and treatment (7), pointing towards favorable outcomes for early treated cbl-responsive MMA patients in previous reports (10). However, also for cbl-nonresponsive MMA patients improved neurological outcomes were reported in one national and one large European cohort after identification by NBS and early treatment compared to the natural history cohort (13, 31). In the present study, all individuals with mut^0^-type MMA experienced a symptomatic disease course with impaired fine motor skills reported in a quarter and impaired gross motor skills reported in half of the patients.

According to previous studies, mortality rate in screened PA remains high (20-30%) despite early detection and guideline concordant treatment (14, 31). Therefore, the benefit of NBS for individuals with PA is less evident: Although NBS allows earlier diagnosis and treatment it does not prevent metabolic decompensations and frequent hospital admissions (14). Of note, more than half (7/13) of the individuals with PA experienced an EO decompensation, four already before the NBS result was available. This is consistent with previous studies reporting at least half of the PA patients to be symptomatic at the time of the NBS result (13, 14, 31). These data highlight that NBS leads to earlier diagnosis especially in LO patients (13). Focusing on morbidity, our study with a limited observation period found that over 80% of the screened individuals with PA were symptomatic, with neurological symptoms being particularly common. Compared to symptomatically diagnosed PA patients, NBS has previously not be associated with improved neurological outcomes (13, 14). Similar results were found for cognitive outcome: the mean IQ of 90 in our screening cohort seems to be a bit better than in previous studies which showed that three quarters of PA patients were at least mildly retarded (IQ below 70) and that the IQ negatively correlated with the number of metabolic decompensation reported (14). Subsequently, less than half of the children with PA attend regular kindergarten and schools but schools for special needs despite NBS and subsequent treatment (14). In contrast, identification by NBS may reduce or delay cardiac symptoms: individuals with PA identified by NBS have been shown to have a lower risk of developing progressive heart disease compared to those identified after the occurrence of first symptoms (13). In our young age cohort, 30% of the screened PA patients presented with cardiac symptoms by the last visit: one child with cardiomyopathy, and three children with prolonged QT_c_ intervals.

### Attenuation of the clinical phenotype in cblC deficiency

Individuals with cblC deficiency were previously reported to benefit from early metabolic treatment, improving survival and reducing or preventing failure to thrive, hematological abnormalities, hemolytic uremic syndrome, and hydrocephalus (19, 21, 22, 32). However, the effect of early therapy on the ocular complications, i.e. optic nerve atrophy, was limited (19, 21, 22, 32). In the present study, all screened individuals with cblC deficiency experienced a symptomatic disease course, 80% were already symptomatic at the time of diagnosis and treatment initiation (e.g feeding difficulties, nystagmus). One child who was initially asymptomatic, developed first symptoms (nystagmus and optic atrophy) in pre-school age and presented with optic atrophy but no further neurological symptoms at last follow-up in adolescence. This is in line with previous reports on early diagnosed and treated individuals with cblC deficiency presenting with less severe disease courses similar to those with attenuated cblC deficiency (19, 33). In the present study, optic nerve atrophy was reported in three out of five individuals until the last visit, while hemolytic uremic syndrome or hydrocephalus were not found. The reported impact of early postnatal treatment on the neurocognitive outcome of patients with cblC deficiency is inconsistent (19, 21, 22, 32), while normal cognitive development was reported for one child with prenatal start of treatment (34). Similarly, DDST and IQ test results varied in our cohort, ranging from normal development to learning disability (IQ 79) or severe developmental delay. None of them attended regular kindergarten, and all but one attended a school for special needs or received other support.

### Clinical benefit for individuals with homocystinurias (CBS and MTHFR deficiency)

In the natural disease course of MTHFR deficiency, first symptoms occur at a median age of 1-2 months (20, 21, 35). Children identified by family screening prior to the onset of first symptoms (age at diagnosis not reported in detail (20)) showed a reduction in the severity of the clinical course, i.e. a normal cognitive outcome compared with symptomatically diagnosed patients (median age at diagnosis 32 days; range 6-90 days) (20, 36). These results are also confirmed by the presented study. Two children with MTHFR deficiency were identified pre-symptomatically and treated according to a recent guideline. They developed normally, did not develop disease-related symptoms until the last follow-up visit (3.5 years visit), and attended a regular kindergarten.

We identified only one individual with CBS deficiency (full responsive to pyridoxine) (7). Unlike individuals with non-responsive (classic) or partially responsive forms of CBS deficiency, individuals with full responsiveness to pyridoxine have so far been assumed to be missed by NBS due to mild metabolic derangement (5, 15, 18, 37). NBS allows to identify individuals with CBS deficiency long before the onset of first clinical symptoms at a median of 4 years in classic CBS deficiency and 7 to 13 years of life in pyridoxine-responsive CBS deficiency (15, 18, 35). Early treatment often prevents the onset of severe symptoms and complications in CBS deficiency (16, 17), especially ophthalmologic symptoms (e. g. lens luxation) and thromboembolic events, further the neurologic symptoms and developmental disability are at least evidently mitigated, which is especially true in the attenuated cofactor-responsive disease variants (15, 18, 19, 21). The child with full responsiveness to pyridoxine remained asymptomatic until the last follow-up visit (7). Nonresponsive patients tend to experience more severe symptoms (17), while early treatment in infancy leads to improved clinical outcomes without major disease-related complications (16).

### Neonatal cobalamin deficiency

In line with a previous report (29), we confirmed that individuals with neonatal cbl deficiency identified by NBS and early treated have excellent clinical and cognitive outcomes.

### Limitations

One limitation of this study is the relatively small cohort size, an inherent challenge in all studies on individuals with rare diseases. With a median follow-up of 3.6 years the study provides only a medium-long observational period and therefore cannot report on disease-specific complications during adolescence or adulthood. Further studies in larger international screening cohorts are desirable.

## Conclusions

While early diagnosis through NBS is possible and together with early treatment according to current guidelines improves the clinical outcome in individuals of the NBS pilot cohort, disease-specific differences should be carefully considered. In general, the greatest benefit is seen in those who do not develop first symptoms before NBS results are available and in diseases with high treatment efficacy. Therefore, the greatest benefit has been seen for individuals with neonatal cbl deficiency, cbl-responsive MMA as well as cblC, MTHFR and CBS deficiency, while the benefit of NBS is less evident for individuals with cbl-nonresponsive MMA and PA.

## Synopsis

Early detection through NBS and subsequent specialized metabolic care improve clinical outcomes and survival in CBS and MTHFR deficiency, and to some extent in cblC and cobalamin-responsive MMA while the benefit for individuals with PA and cobalamin-nonresponsive MMA is less evident due to the high (neonatal) decompensation rate, mortality, and long-term complications.

## Details of the contributions of individual authors

Dr Mütze, Prof Hoffmann, and Prof Kölker conceptualized and designed the study and the data collection instruments. Dr Mütze und Prof Kölker coordinated and supervised data collection, evaluated and interpreted data, and drafted the manuscript. Dr Reischl-Hajiabadi co-designed the study, collected NBS and clinical data, carried out statistical analyses, evaluated and interpreted data and drafted the initial manuscript. Drs Schnabel, Mengler, and Posset collected NBS and clinical data and critically reviewed and revised the manuscript for important intellectual content. Mr Gleich carried out data collection and management, and critically reviewed and revised the manuscript. Drs Lommer-Steinhoff and Burgard carried out psychological testing, collected clinical data and critically reviewed and revised the manuscript. Profs Gramer, Grünert, Hennermann and Freisinger and Drs Lindner, Thimm and Krämer collected the comprehensive follow-up data at their study sites and reviewed and critically revised the manuscript. Drs Hörster, Lenz and Christ examined patients, collected data and revised the manuscript. Dr Garbade carried out and supervised statistical analyses and critically reviewed and revised the manuscript. The corresponding author (Dr Mütze) had full access to the complete dataset of the study and had final responsibility for the decision to submit for publication. All authors approved the final manuscript as submitted and agree to be accountable for all aspects of the work.

## Corresponding Author

PD Dr. med. Ulrike Mütze

## Details of ethics approval

The NBS Pilot study was approved by the local ethics committee (University Hospital Heidelberg, applications number S-533/2015). The NBS 2025 outcome study was first approved by the local ethics committee of the coordinating site (University Hospital Heidelberg, applications number S-104/2005) and then by the contributing sites.

## Patient consent statement

All procedures followed were in accordance with the ethical standards of the responsible committee on human experimentation (institutional and national) and with the Helsinki Declaration of 1975, as revised in 2013. Informed consent was obtained from all patients and/or caregivers for being included in the study.

## Data sharing statement

Deidentified individual participant data will not be made available.

## Abbreviations

C3: propionyl carnitine
cbl: cobalamin
cblC: cobalamin C
CBS: cystathionine-β-synthase
DDST: Denver Developmental Screening Test
DESTATIS: Federal Statistical Office
DGKJ: German Society for Pediatrics and Adolescent Medicine
EO: early onset
IMDs: inherited metabolic diseases
LO: late onset
M: biochemical checkups as well as clinical monitoring for comorbidities
MMA: methylmalonic acidemia
MTHFR: methylenetetrahydrofolate reductase
mut: methymalonyl-CoA mutase
NBS: newborn screening
OA: organic acidurias
PA: propionic acidemia
T: therapy recommendations

## Data Availability

All data produced in the present work are contained in the manuscript

