## Supplementary Table for "Clinical outcomes and survival of individuals with methylmalonic acidemia, propionic acidemia, classic homocystinuria, and remethylation disorders identified through newborn screening"

### Supplementary Material

**Table: Disease-related symptoms** Disease-specific symptoms for each disorder are highlighted in gray. *Possible onset under therapy with vitamine $B_6$. **After dislocation or lens removal.

| **Disease-related symptoms** | **PA** | **mut$^{0}$-type MMA** | **cblA-type MMA** | **CBS deficiency** | **MTHFR deficiency** | **cblC deficiency** |
|---|---|---|---|---|---|---|
| Episodes of coma/somnolence/acute neurologic event since last presentation? | | | | | | |
| Impaired cognition | | | | | | |
| Speech delay (comprehension/processing) | | | | | | |
| Speech delay (articulation/motor function) | | | | | | |
| Impaired fine motor skills | | | | | | |
| Impaired gross motor skills | | | | | | |
| Behavioral problems | | | | | | |
| Ataxia | | | | | | |
| Chorea | | | | | | |
| Dystonia | | | | | | |
| Tremor (resting/holding) | | | | | | |
| Myoclonia | | | | | | |
| Signs of brainstem involvement | | | | | | |
| Sensory impairment | | | | * | | |
| Epilepsy | | | | | | |
| Loss of functions | | | | | | |
| Exercise intolerance | | | | | | |
| Muscle weakness | | | | | | |
| Muscular hypotonia | | | | | | |
| Muscular hypertonia/ spasticity | | | | | | |
| Hyperreflexia (PSR) | | | | | | |
| Hyporeflexia (PSR) | | | | * | | |
| Scoliosis | | | | | | |
| Cardiomyopathy | | | | | | |
| Prolonged QTc interval | | | | | | |
| Myopia | | | | | | |
| Nystagmus | | | | | | |
| Aphakia | | | | ** | | |
| Pseudophakia | | | | | | |
| Lens dislocation | | | | | | |
| Retinal pigmentary degeneration | | | | | | |
| Optic atrophy | | | | | | |
| Visual field defects | | | | | | |
| Hearing impairment | | | | | | |
| Hepatomegaly | | | | | | |
| Acute liver failure | | | | | | |
| Chronic liver failure | | | | | | |
| Elevation of transaminases | | | | | | |

| |
|---|
| Liver transplant |
| Tubulopathy |
| Renal insufficiency |
| Kidney transplant |
| Pancytopenia |
| Neutropenia |
| Anemia |
| Hair abnormalities |
| Skin abnormalities |
| Weight <3. Perc. |
| Length <3. Perc. |
| Length >97. Perc. |
| Feeding difficulties |
